# Comparison of Early and Intermediate-Term Outcomes Between Hybrid Arch Debranching and Total Arch Replacement: A Systematic Review and Meta-analysis of Propensity-Matched Studies

**DOI:** 10.1101/2024.11.12.24317156

**Authors:** Naritsaret Kaewboonlert, Worawong Slisatkorn, Apichat Tantraworasin, Punthiti Pleehachinda, Tossapol Prapassaro, Natthipong Pongsuwan, Chanut Chatkaewpaisal, Tummarat Ruangpratyakul

## Abstract

**OBJECTIVES:** To systematically review and pool the clinical outcomes of hybrid arch repair (HAR) and total arch replacement (TAR) with or without a frozen elephant trunk for treating aortic arch aneurysms, dissections, or other pathology in propensity score-matched studies.

**METHODS:** We conducted electronic database searches in PubMed, Embase, the Cochrane Library, and Google Scholar to identify studies reporting outcomes of HAR versus TAR. Risk of bias was assessed using non-randomized studies of interventions (ROBINS-I) tool. The primary outcome was in-hospital mortality analyzed using a random-effects model to compute the odds ratio (OR). Survival probability was expressed as hazard ratios (HR) calculated through the inverse variance method. The results were reported following the Preferred Reporting Items for Systematic Reviews and Meta-Analyses (PRISMA) guidelines.

**RESULTS:** This meta-analysis included 13 studies with 3,392 patients. There was no significant difference in in-hospital mortality between HAR and TAR groups (OR 1.08; 95% CI 0.78-1.49; p = 0.630). However, HAR group showed a higher incidence of permanent neurological dysfunction (PND) (OR 1.71; 95% CI 1.22-2.41; p < 0.001). In subgroup meta-analysis with isolated type A aortic dissection (ITAAD), HAR showed significantly lower in-hospital mortality (p = 0.040) but no difference in PND. Other post-operative complications were significantly lower in the HAR group for renal failure (OR 0.65; 95% CI 0.49-0.87; p < 0.001), sternal re-entry due to bleeding (OR 0.55; 95% CI 0.34-0.89; p = 0.010), and tracheostomy (OR 0.61; 95% CI 0.38-0.96; p = 0.030). There is no statistical difference in 3-year survival probability (HR 0.97; 95% CI 0.70-1.35; p = 0.870).

**CONCLUSIONS:** TAR has more favorable than HAR in MDAD patients, offering lower rates of neurological dysfunction and better 3-year freedom from re-intervention. For ITAAD patients, HAR potentially provides better in-hospital mortality and 3-year survival rates, with fewer complications such as renal failure, re-sternotomy, and tracheostomy.

## INTRODUCTION

The most common aortic arch pathology is atherosclerosis, followed by other conditions such as aortic dissection(1, 2). Total arch replacement (TAR) is an aggressive and high-risk surgical procedure. The aim of the surgical treatment for chronic aortic aneurysms is to prevent aortic rupture or dissection and to reduce the incidence of serious postoperative complications such as death, permanent neurological dysfunction, acute renal failure, and tracheostomy. Moreover, the procedure assigned to the patients should effectively improve the long-term survival probability and reduce the risk of disability(3, 4).

Due to postoperative morbidity and mortality, less invasive treatment options such as thoracic endovascular repair (TEVAR) with aortic arch surgical debranching or hybrid arch repair (HAR)(3) are introduced for selected patients who are elderly or not fit for TAR.

In 2024, the European Association for Cardio-Thoracic Surgery (EACTS) and the Society of Thoracic Surgeons (STS) guideline(5), there is an increasing trend for adopting TEVAR with HAR as an option in complex arch aneurysms or acute type A aortic dissections(6-8). The risks and benefits of HAR compared to TAR for the treatment of TAAD remain under debate(8-10).

Currently, TAR is the standard treatment for patients with aortic arch aneurysms or TAAD. The individualized selection strategy for surgical treatment options remains controversial due to a lack of solid data comparing various strategies(5). There have been no randomized controlled trials comparing the outcomes of different treatments. Most studies on the outcomes of HAR versus TAR are based on observational data.

Therefore, we conducted a meta-analysis to compare the early clinical outcomes and intermediate-term survival of patients with aortic arch aneurysms or TAAD undergoing TAR or HAR. The included studies were matched by propensity scores in order to reduce selection bias.

## METHODS

Systematic review was conducted by accordance with the Preferred Reporting Items for Systematic Reviews and Meta-Analysis (PRISMA) guidelines outlined by Page et al.(11).

### Population, Intervention, Comparison, and outcome (PICO)

Using the PICO framework, as described by Richardson et al.(12), the research question was addressed through this analysis as follows:

- **Population:** This includes any patients diagnosed with an aortic arch aneurysm or acute and chronic aortic dissections involving the aortic arch that required repair. The repair method used was debranching combined with thoracic endovascular aortic repair. These patients were subsequently matched with similar patient characteristics within the studies to those who had undergone conventional total arch replacement (in retrospective cohort studies where propensity score matching occurred).
- **Intervention:** any patients indicated for hybrid arch debranching repair.
- **Comparison:** any patients indicated for total arch replacement whose characteristics matched those of the intervention group.
- **Outcomes:** outcome measures were divided into:

- *Early outcome measures:* These included in-hospital mortality, permanent neurological dysfunction or stroke, renal failure, sternal re-entry due to bleeding, and tracheostomy, all considered as odds ratio (OR).
- *Intermediate term outcome measure:* These included 3-year and 5-year survival probability, as well as 3-year and 5-year freedom from re-intervention as hazard ratio (HR)

### Search strategy

In April 2024, we conducted a search for studies related to this topic across multiple databases, including PubMed, Embase, Cochrane Library, and Google Scholar. We also performed a manual search for references from published studies that met the criteria of this study. The search encompassed all fields under the following headings: "hybrid arch" AND "total arch" AND "outcome" AND "aneurysm," using medical subject headings (MeSH Terms). These terms were connected by the Boolean operator ‘AND’. The search was not limited by language or publication year. Initially, the titles of retrieved studies were screened, followed by a thorough evaluation of the study abstracts and full texts to identify studies suitable for inclusion.

### Eligible criteria

Studies were considered eligible if they met the following inclusion criteria: (1) studies to be prospective observation or retrospective cohort with propensity-matched design, (2) studies to be comparison of outcome in adult patients aged more than 18 years, (3) studies had to reported for early and intermediate outcomes. Studies were excluded if satisfied any one of the following exclusion criteria: (1) studies where participants were not propensity score matched, (2) case report, correspondence, perspective or review article, (3) no arch involvement, and (4) thoracoabdominal aortic aneurysm involvement.

### Data extraction and quality assessment

The literature search was conducted by two independent reviewers (NK and NP) using pre-designed search strategies. Duplicate studies were manually removed. Each reviewer systematically screened titles, abstracts, and, where necessary, full texts to identify studies that met the inclusion criteria. Data from the retrieved manuscripts—including study information, design, patient demographics, treatment details, and early and intermediate outcomes—were extracted. The risk of bias was assessed according to the Cochrane Handbook for Systematic Reviews of Interventions(13), using the Risk of Bias in Non-randomized Studies of Interventions (ROBINS-I) tool(14).

### Statistical analysis

The data reported as medians with interquartile ranges were estimated using the Hozo et al. approximation(15). Age was pooled using a fixed effects model to estimate the mean weighted in both groups. Descriptive statistics were primarily used to determine associations in patient characteristics between HAR and TAR using exact tests and independent t-tests, as appropriate. Early surgical outcomes were expressed as odds ratios (OR) for binary outcomes, and effect sizes for two-group comparisons were computed using a random effects model with the restricted maximum-likelihood method. The outcome of different surgical option on survival outcomes and freedom from re-intervention were measure as hazard ratio (HR), if HR not available, the data were extracted from Kaplan-Meier curve using WebPlotDigitizer (available from: https://automeris.io/WebPlotDigitizer) then calculate the HR with 95% confidence interval (CI) using survival probabilities at difference time intervals from the survival curve and the number of patients at-risk in each time intervals. The HR was transformed to natural logarithm before aggregating by inverse variance method, then expressed as the HR with 95%CI in the forest plot(16-18). Subgroup meta-analyses were performed in order to explore causes of heterogeneity. Subgroups were categorized based on the study population domain, including studies reporting outcomes for isolated type A aortic dissections (ITAAD) and mixed degeneration and dissection (MDAD) arch pathology. The symmetry in funnel plots was used to evaluate publication bias. Leave-one-out sensitivity analysis was used to assess robustness of the synthesized results. All tests were two-tailed, with a significance level set at p < 0.05. Descriptive statistics, meta-analysis, and chart creation were facilitated by STATA statistical software, version 17 (Stata Corp, College Station, Texas) and Microsoft Excel (Microsoft Corp., Redmond, Washington).

## RESULTS

### Literature search and study characteristics

The study selection process is outlined in Figure 1. A total of 387 articles were identified, with 97 records removed due to duplication and 266 excluded after screening. Additionally, 28 records were excluded for not meeting the eligibility criteria: 19 were non-matching studies, 5 were correspondence, review articles, or case reports, 3 did not involve the aortic arch, and 1 involved a thoracoabdominal aortic aneurysm. The study characteristics and risk of bias assessment are detailed in Table 1. All of the included studies are retrospective cohort design and adjust the confounding factors using propensity score matching method. There is no one studies reporting the missing data in the cohort; these are risks for reporting the effect size of missing data. Seven of thirteen studies(10, 19-24) presented a serious risk of bias, as they did not incorporate known confounding factors, such as cerebrovascular disease or chronic renal failure, into the propensity model (Table 1). Two of thirteen studies(25, 26) showed a serious risk of bias due to risk of deviations from intended interventions and the classification of interventions. We rated nine out of thirteen studies(10, 19-26) as having an overall serious risk of bias, while the remaining studies(27-30) demonstrated a moderate overall risk. None of the studies had an overall low risk of bias (Supplementary Figure 1), highlighting the inherent limitations of non-randomized studies.

**Figure 1.**
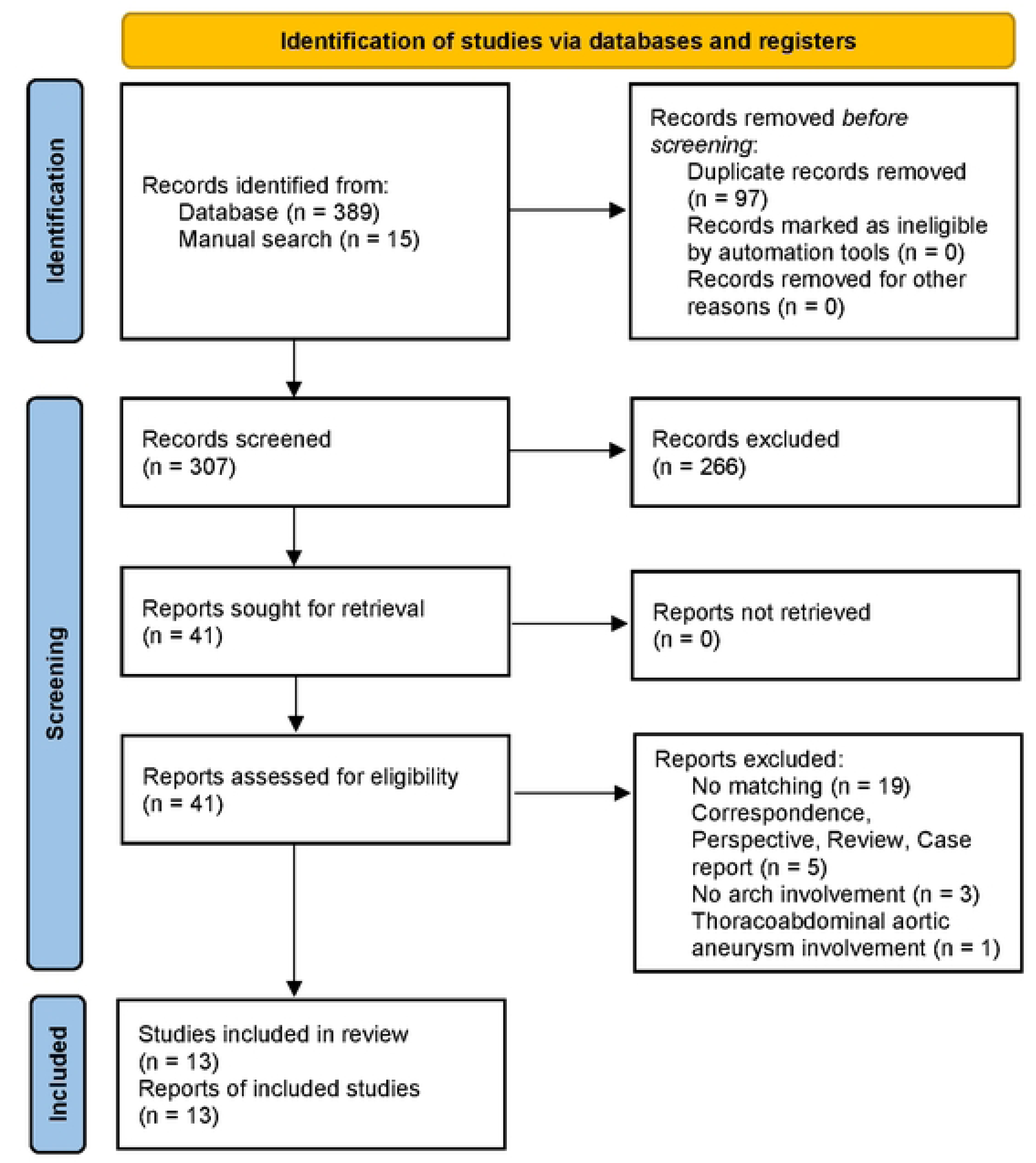
PRISMA flow diagram shows the systematic review process.

**Table 1.**
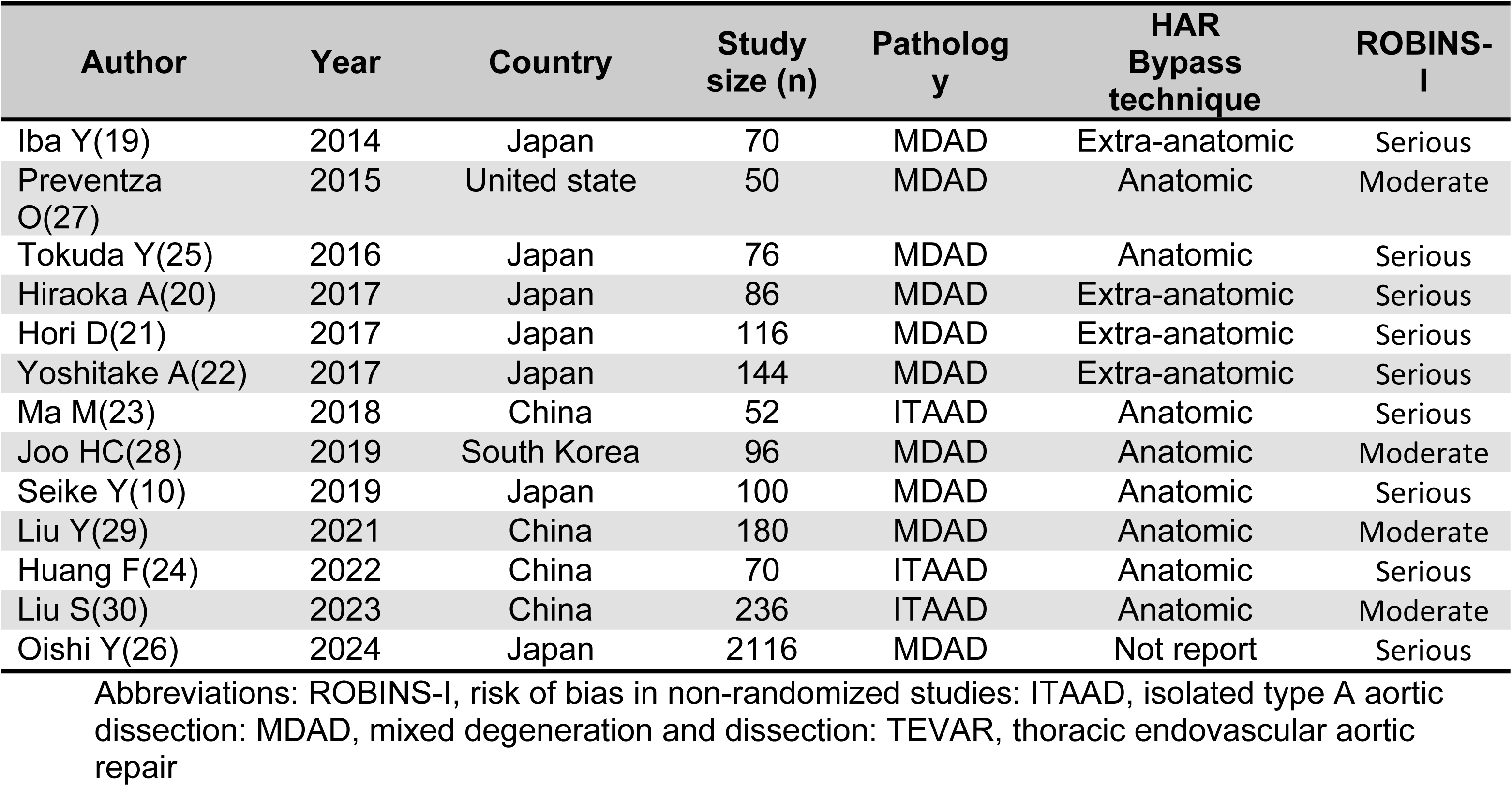
Characteristics of studies included in this systematic review with risk of bias assessment.

### Patient characteristics

A total number of 3,392 patients was included in the studies, with 1,696 patients in each group, matched using the propensity score model (Table 2). The mean age for the HAR and TAR group were 74.67 ± 14.17 and 74.38 ± 13.05, respectively. There was no statistical difference between the means of ages both groups (p = 0.535). In the present study, the percentage of males in the HAR group (75.9%) and the TAR group (75.2%) showed no statistically significant difference (p = 0.689). The proportion of urgent and emergency operations was 27.2% in the HAR group and 29.8% in the TAR group, with no statistically significant difference (p = 0.363). There was no statistically significant difference in the prevalence of underlying diseases, such as hypertension, diabetes mellitus (DM), coronary artery disease (CAD), previous myocardial infarction (MI), chronic obstructive pulmonary disease (COPD), and renal failure.

**Table 2.** Summary of clinical characteristics reported in the included studies, presented as mean ± standard deviation or frequency and percentage.

| Characteristics | Number of patients | HAR | Number of patients | TAR | p-value |
| --- | --- | --- | --- | --- | --- |
| Age, year | 1696 | 74.67±14.17 | 1696 | 74.38±13.05 | 0.535 |
| Male gender | 1696 | 1287 (75.9) | 1696 | 1276 (75.2) | 0.689 |
| Hypertension | 1536 | 1261 (82.1) | 1536 | 1274 (82.9) | 0.569 |
| Diabetic mellitus | 1536 | 275 (17.9) | 1536 | 274 (17.8) | 1.000 |
| Coronary artery disease or previous myocardial infarction | 404 | 114 (28.2) | 404 | 104 (25.7) | 0.476 |
| Chronic obstructive pulmonary disease | 1638 | 178 (10.9) | 1638 | 172 (10.5) | 0.822 |
| Renal failure | 1603 | 181 (11.3) | 1603 | 162 (10.1) | 0.304 |
| Urgent or emergency operation | 580 | 158 (27.2) | 580 | 173 (29.8) | 0.363 |

Abbreviations: HAR, hybrid arch repair; TAR, total arch replacement

### Early outcome

#### Early mortality analysis

All 12 studies were included in a meta-analysis of in-in hospital mortality. Forrest plots were calculated and revealed no heterogeneity (I^2^ = 0%). The overall data showed no significant difference in in-hospital mortality between HAR and TAR groups (OR 1.08; 95% Cl 0.78-1.49; p = 0.630) (Figure 2). In the MDAD and ITAAD subgroups meta-analysis showed no significant difference in in-hospital mortality between the two groups ((OR 1.20; 95% CI 0.86-1.68) and (OR 0.36; 95% CI 0.12-1.07) respectively). A test for subgroup differences revealed a statistically significant difference between the two groups (p = 0.040) (Figure 2). Publication bias was assessed using a funnel plot (Supplementary Figure 1) along with regression-based Egger’s test (p = 0.653) and Begg’s test (p = 0.631), both indicating no significant publication bias. The leave-one-out sensitivity analysis confirmed that the results were robust and not significantly affected by the exclusion of any single study (Supplementary Figure 2).

**Figure 2.**
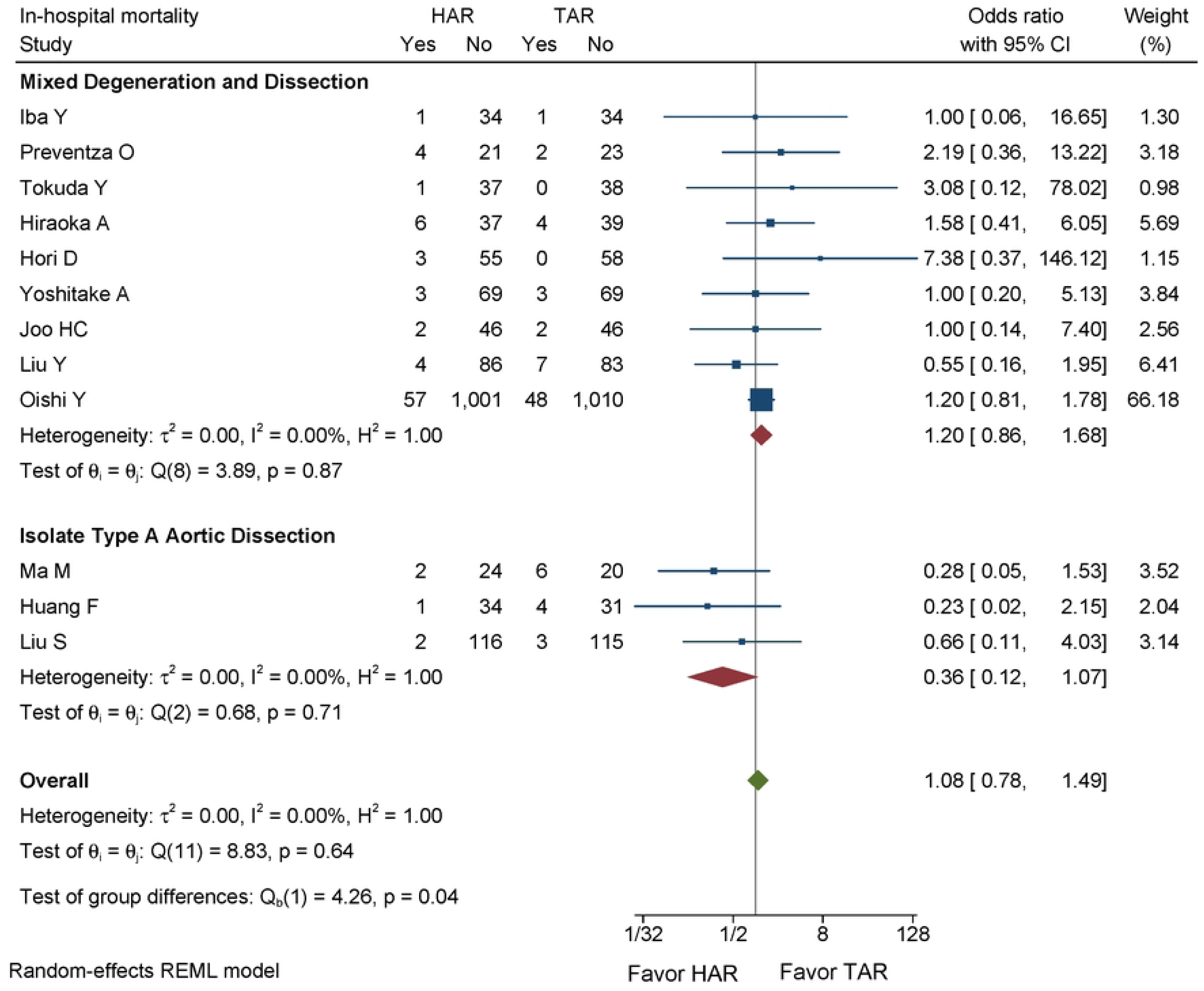
Forest plot showing the results of in-hospital mortality after HAR versus TAR with the subgroup meta-analysis in ITAAD and MDAD aortic arch pathologies. HAR: Hybrid Arch Repair; TAR: Total Arch Replacement; ITAAD: Isolated Type A Aortic Dissection; MDAD: Mixed Degeneration And Dissection.

#### Permanent neurological dysfunction and other post-operative complications

The incidence on permanent neurological dysfunction (PND) or stroke showed a higher trend in patients undergoing HAR compared to those in the TAR group (OR 1.71; 95% CI 1.22-2.41; p <0.001; I^2^ = 0%). However, the subgroup meta-analysis for ITAAD studies found no statistically significant difference in this outcome (Figure 3).

**Figure 3.**
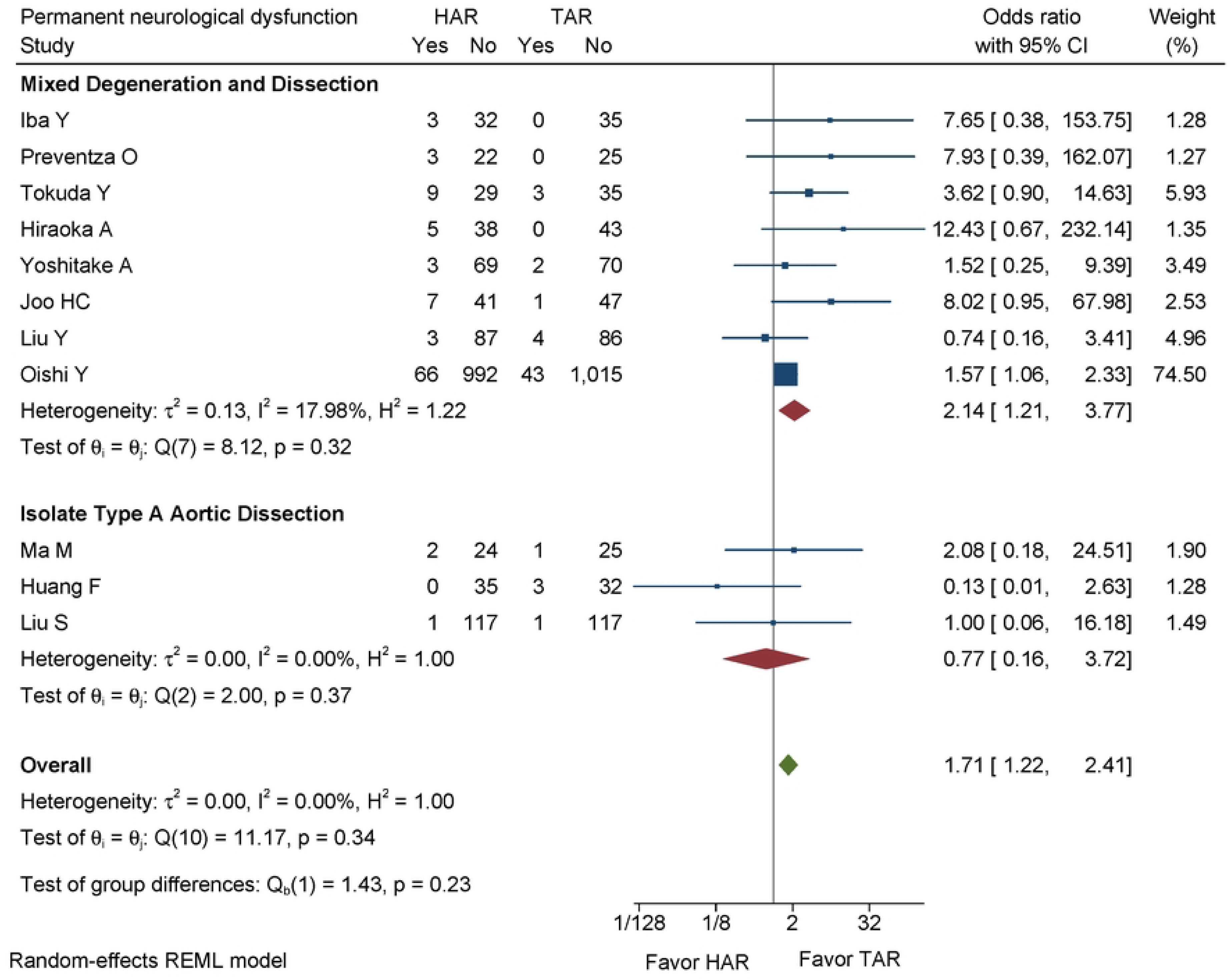
Forest plot showing the results of permanent neurological dysfunction after HAR versus TAR with the subgroup meta-analysis in ITAAD and MDAD aortic arch pathologies. PND: Permanent Neurological Dysfunction; HAR: Hybrid Arch Repair; TAR: Total Arch Replacement; ITAAD: Isolated Type A Aortic Dissection; MDAD: Mixed Degeneration And Dissection.

Other pooled post-operative complication results were found, a lower occurrence of renal failure in the HAR group compared to the TAR group (OR 0.65; 95% CI 0.49-0.87; p < 0.001; I^2^ = 0%) (Supplementary Figure 3). A lower occurrence of sternal re-entry due to bleeding was observed in the HAR group compared to the TAR group (OR 0.55; 95% CI 0.34-0.89; p = 0.010, I^2^ = 0%) (Supplementary Figure 4). Similarly, a lower occurrence of tracheostomy was observed in the HAR group compared to the TAR group (OR 0.61; 95% CI 0.38-0.96; p = 0.030; I² = 6.80%) (Supplementary Figure 5).

### Intermediate term survival rate

Ten studies were included(10, 19-23, 27-30), comprising 1130 patients, stratified as 565 pairs-matched in each group. The pooled results for the 3-year survival probability showed no statistical difference between the HAR and TAR groups (HR 0.97; 95% CI 0.7-1.35, p = 0.870, I^2^ = 46.75%) (Figure 4). In the ITAAD subgroup, the 3-year survival probability was more favorable for HAR (HR 0.36; 95% CI 0.16-0.79; I^2^ = 7.52%). Conversely, in the MDAD subgroup, there was no statistically significant difference in the hazard ratio between the HAR and TAR groups (HR 1.21; 95% CI 0.84-1.73; I^2^ = 15.06%) (Figure 4). The test for subgroup differences indicates a statistically significant result (p = 0.010), suggesting that type A aortic dissection is significantly modified the effect of HAR compared to TAR.

**Figure 4.**
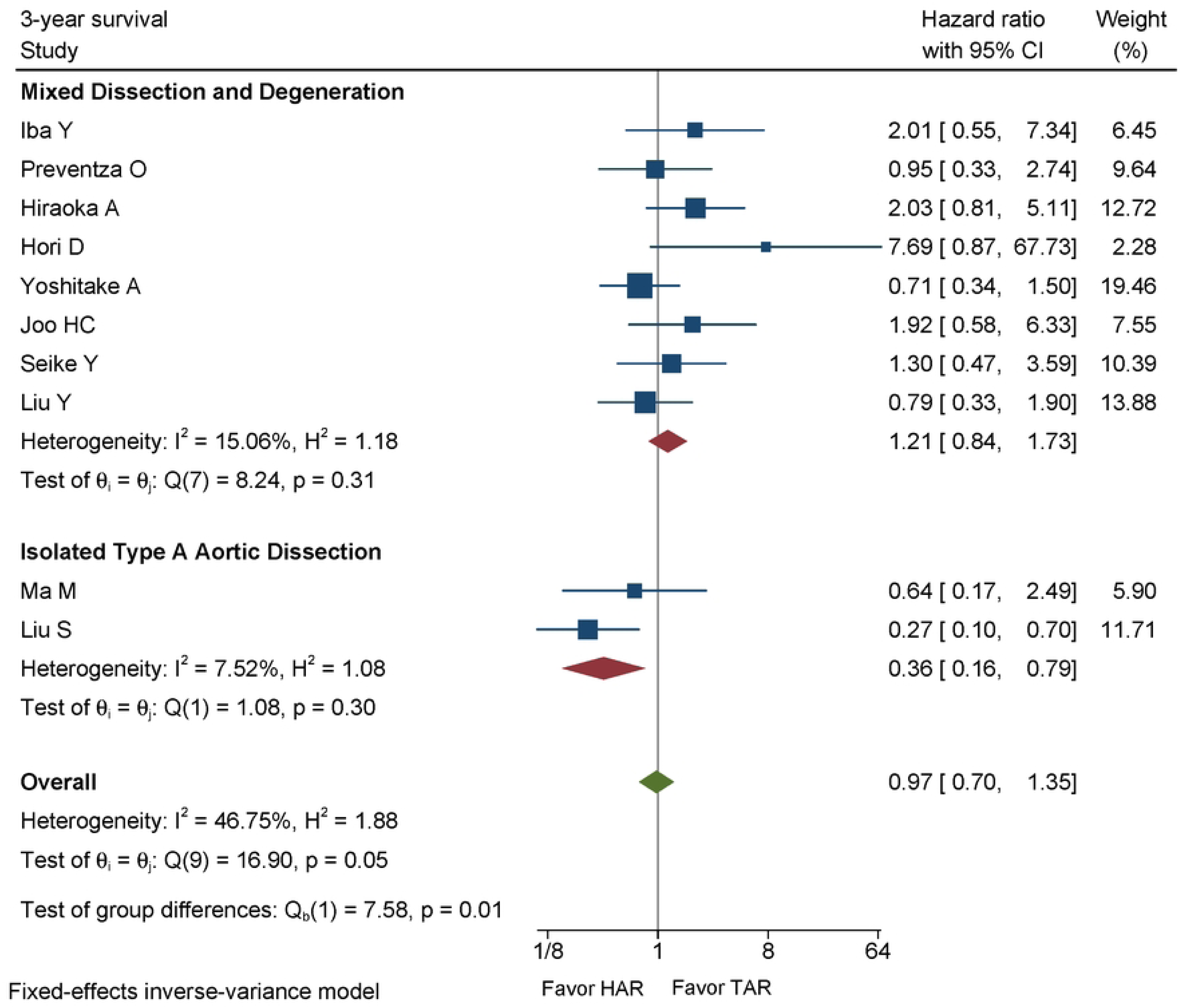
Forest plot showing the results of 3-year survival probability after HAR versus TAR with the subgroup meta-analysis in ITAAD and MDAD aortic arch pathologies. HAR: Hybrid Arch Repair; TAR: Total Arch Replacement; ITAAD: Isolated Type A Aortic Dissection; MDAD: Mixed Degeneration And Dissection.

Only four studies(10, 21, 22, 28) reported the 5-year survival outcome. The pooled result showed a more favorable outcome in the TAR group compared to the HAR group, but this difference was not statistically significant. (HR 1.29; 95% CI 0.81-2.05; p = 0.280; I^2^ = 53.25%) (Supplementary Figure 6).

### Freedom from re-intervention probability

Five studies(19, 21, 22, 28, 29) were included, comprising 606 patients stratified as 303 pair-matched in each group. The overall result for the 3-year freedom from re-intervention indicated a significantly higher rate of re-intervention in the HAR group compared to the TAR group (HR 3.69; 95% CI 1.97-6.90; p < 0.001; I^2^ = 31.16%) (Figure 5). At the 5-year follow-up, three studies(21, 22, 28) reported outcomes, also showing a higher rate of re-intervention in the HAR group (HR 4.39; 95% CI 2.31-8.34; p < 0.001; I^2^ = 0%), with no heterogeneity among the studies (Supplementary Figure 7).

**Figure 5.**
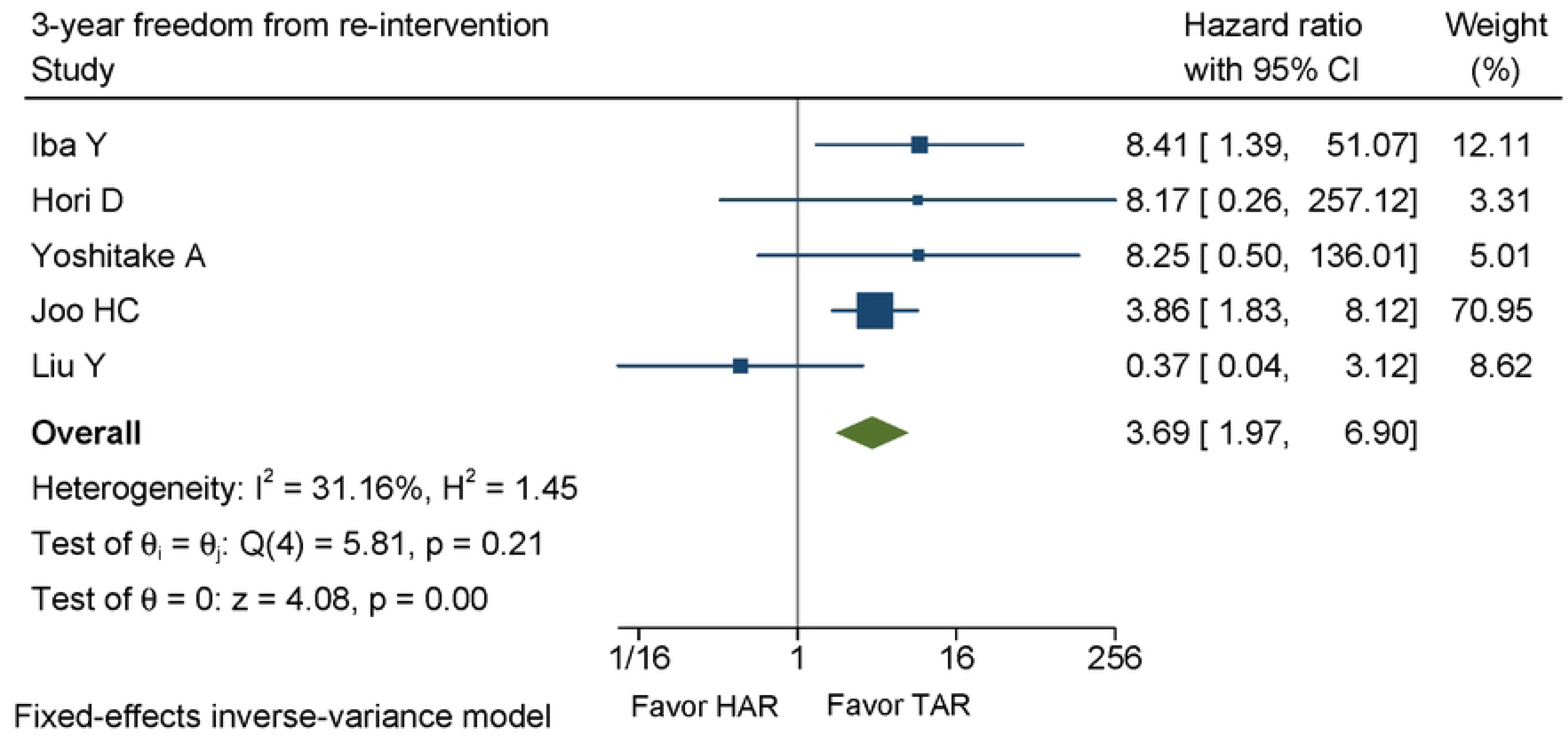
Forest plot showing the results of 3-year freedom from re-intervention after HAR versus TAR. HAR: Hybrid Arch Repair; TAR: Total Arch Replacement.

## DISCUSSION

The primary outcome of our analysis indicated no overall difference in in-hospital mortality across all studies. However, subgroup analysis revealed a statistically significant difference between the overall in-hospital mortality and subgroup involving ITAAD studies (p = 0.040). In fact, our data suggested some pattern that could have been indicative of trend in-hospital mortality benefit among ITAAD patients undergoing HAR compared to TAR (OR 0.36; 95% CI 0.12-1.07). This finding was consistent with previous studies(23, 24, 30), on patients with ITAAD undergoing Type II hybrid debranching, as proposed by Bavaria et al. (31). The propensity-matched cohort studies showed that in-hospital mortality ranged from 1.7% and 7.7% in the patients with HAR and 2.5% to 23.1% in patients with TAR(23, 24, 30), regardless of whether they had undergone a frozen elephant trunk procedure. In our study results, in-hospital mortality is comparable to the study reported by Bavaria et al. for Type I and Type II HAR in patients with aortic arch aneurysms(31).

Our analysis estimated the odds ratio of PND following HAR versus TAR. In studies involving MDAD aortic pathologies, there was a significantly higher occurrence of stroke in the HAR group compared to the TAR group (OR 1.71; 95% CI 1.22-2.41; p < 0.001). This indicates a statistical significant difference in stroke occurrence between the two groups. This finding was consistent with literature from studies by Iba et al. (19) and Hiroaka et al.(20), which also reported higher risk of stroke in the HAR. However, no significant difference was found in the subgroup of study involving ITAAD. In contrast, Eleshra et al.(32) reported stroke rates of 14% in patients with degenerative aneurysms and 2% in patients with aortic dissection. Huang F et al.(24) reported a lower occurrence of stroke in HAR among patients with ITAAD, although these findings were not statistically significant (OR 0.13; 95% CI 0.01-2.63).

Our study also revealed a statistically significant reduction in the occurrence of renal failure (OR 0.65; 95% CI 0.49-0.87; p < 0.001), sternal re-entry due to bleeding (OR 0.55; 95% CI 0.34-0.89), and tracheostomy (OR 0.61; 95% CI 0.38-0.96). These results aligned with findings from most other studies(21, 26-28). A large non-propensity matched cohort study by Wallen et al.(33) reported lower rate of mortality, stroke, paralysis, and renal failure rates in the TAR group, this highlighting the potential influence of selection bias, as the TAR group tended to include younger patients with fewer cases of peripheral arterial disease, and less preoperative hemodialysis.

We reported a pooled 3-year survival probability between HAR and TAR, with no statistically significant difference (HR 0.97; 95% CI 0.70-1.35). However, significant differences were observed between the overall result and the ITAAD subgroup. These meta-analyses also suggested a trend that ITAAD may benefit from HAR, but further evidence is needed to support this hypothesis.

The rate of re-intervention at 3 years was significantly higher in the HAR group (HR 3.69; 95% CI 1.97-6.89, p < 0.001), suggesting inferior intermediate outcomes compared to TAR in terms of freedom from re-intervention. However, in the subgroup of ITAAD, HAR showed a significantly lower re-intervention rate than TAR. The 5-year survival possibility favored of the TAR group, but the difference was not statistically significant, likely due to the limited number of patients and propensity score–matched studies.

Recently, Spath et al.(34) published a meta-analysis that pooled outcomes of aortic arch repairs involving endovascular techniques for chronic dissections, degenerative aneurysms, penetrating aortic ulcers, and pseudoaneurysms. The overall technical success rate was 95.5%, with an overall 30-day mortality rate of 6.7%. These outcomes may be comparable to those in unmatched cohorts for the HAR group, which reported mortality rates ranging from 2.9% to 11.1%(21, 24, 26-28). The study highlighted that data on immediate to long-term outcomes from total endovascular repairs are limited, raising questions about the durability of endovascular grafts.

The results from prior single-center research may be constrained by several factors including sample size, surgeon experience, and surgical preference, all of which contribute to selection bias, a major confounding factor. These factors may make it challenging to estimate the therapeutic effects of HAR. Although bias can be minimized by using propensity score matching (35), small studies may still lack of statistical power required for demonstrating accurate results(36). The information obtained from this meta-analysis can be useful to determine the therapeutic effects of HAR compared to conventional TAR. The allocation of surgical procedures among patients with various aortic arch pathologies or within distinct subgroups may be guided by these results.

### Implication of this study

Our study is the first systematic review and meta-analysis to estimate treatment outcomes between HAR and TAR, specifically focusing on reducing selection bias by including only propensity-score matched studies. The choice of surgical treatment for aortic arch pathologies remain a crucial part.

The results suggest that for non-Type A Aortic Dissection pathologies, TAR may be more beneficial compared to HAR due to a lower incidence of permanent neurological dysfunction and a higher freedom from re-intervention at 3 years. On the other hand, in patients with Type A Aortic Dissection, HAR may be considered due to the fact that it can reduce in-hospital mortality and offer benefits to improve 3-year survival rates. Additionally, HAR may help reduce the incidence of postoperative complications such as renal failure, re-sternotomy for bleeding, and tracheostomy.

### Limitation

A major limitation of this meta-analysis was incomplete reporting of missing data across the analyzed retrospective cohorts. Therefore, this omission may introduce some biases, leading to an under- or overestimation of the effect size or making it appear falsely precise. Furthermore, the details in the characteristics of aneurysm and the entry site of dissection were not present in enrolled studies. Such details are necessary to assess the applicability of the surgical results to different patient subgroups and may impact the generalizability of our conclusions.

Furthermore, long-term follow-up of surgical interventions, particularly those using propensity score–matched cohorts, is critically needed. Long-term studies are essential to understand the durability and long-term effectiveness of these treatments. Future research should focus on large prospective observational studies, propensity score matching, or randomized controlled trials. These approaches are crucial for obtaining less biased estimates of the effects of surgical interventions and for validating the results of our meta-analysis.

Finally, the development of an algorithm using individual patient data to guide the choice of surgical procedures is essential. This approach aims to optimize the allocation of surgical treatments based on specific clinical characteristics and conditions, maximizing the benefits for patients.

## CONCLUSIONS

This meta-analysis is the first study to critically evaluate and compare the outcomes of HAR and TAR while using propensity score-matched studies, particularly to reduce selection bias. Our findings suggest that TAR may be more favorable than HAR for MDAD studies, as it is associated with lower rates of permanent neurological dysfunction and better 3-year freedom from re-intervention, while showing similar in-hospital mortality and 3-year survival rates. Conversely, HAR appears to offer advantages for ITAAD patients, potentially leading to lower in-hospital mortality, improved 3-year survival, and reduced incidences of renal failure, re-sternotomy due to bleeding, and tracheostomy.

## Data Availability

The data that support the findings of this study can be obtained from the corresponding author upon reasonable request.

## Acknowledgement

We would like to express our sincere gratitude to all the researchers and clinicians who contributed the studies included in this meta-analysis. Their work has been advancing our understanding of the outcome of surgical intervention in different aortic arch pathologies. We also thank the Suranaree University of Technology, Mahidol University, and Chiang Mai University for their support throughout this research process, and the Royal College of Surgeons of Thailand through research funding to present at Lisbon, Portugal. Additionally, we are grateful to our colleagues for their insightful comments and suggestions that helped improve this manuscript.

## Author contributions statement

**Naritsaret Kaewboonlert**: Contributed to all parts of the research. **Punthiti Pleehachinda** and **Tossapol Prapassaro:** Writing, Review and Editing. **Natthipong Pongsuwan:** Conceptualized, Writing, Review and Editing. **Worawong Slisatkorn** and **Apichat Tantraworasin:** Writing, Review and Editing. **Chanut Chatkaewpaisal** and **Tummarat Ruangpratyakul:** Review and Editing.

## Data Available Statement

All data that support the finding of this meta-analysis are available from the corresponding author upon reasonable request.

